# Potential contributors to increased pulmonary embolism hospitalizations during the Covid-19 pandemic

**DOI:** 10.1101/2021.05.18.21257372

**Authors:** Daniela Husser, Sven Hohenstein, Vincent Pellissier, Laura Ueberham, Sebastian König, Gerhard Hindricks, Andreas Meier-Hellmann, Ralf Kuhlen, Andreas Bollmann

**Author notes:** **Corresponding author:** Andreas Bollmann, MD, PhD, Heart Center Leipzig at University of Leipzig, Strümpellstr. 39, 04289 Leipzig, Germany.

## Abstract

**Background:** After the first Covid-19 infection wave, a constant increase of pulmonary embolism (PE) hospitalizations not linked with active PCR-confirmed Covid-19 has been observed but potential contributors to this observation are unclear. Therefore, we analyzed associations between changes in PE hospitalizations and (1) the incidence of non-Covid-19 pneumonia, (2) the use of computed tomography pulmonary angiography (CTPA), (3) volume depletion and (4) preceding Covid-19 infection numbers in Germany.

**Methods:** Claims data of Helios hospitals in Germany were used and consecutive cases with a hospital admission between May 6 and December 15, 2020 (PE surplus period) were analyzed and compared to corresponding periods covering the same weeks in 2016–2019 (control period). We analyzed the number of PE cases in the target period with multivariable Poisson general linear mixed models (GLMM) including (a) cohorts of 2020 versus 2016– 2019, (b) the number of cases with pneumonia, (c) CTPA, and (d) volume depletion and adjusted for age and sex. In order to associate the daily number of PE cases in 2020 with the number of preceding SARS-CoV-2 infections in Germany, we calculated the average number of daily infections (divided by 10,000) occurring 14 up to 90 days with increasing window sizes before PE cases and modelled the data with Poisson regression.

**Results:** There were 2,404 PE hospitalizations between May 6 and December 15, 2020 as opposed to 2,112 – 2,236 (total 8,717) in the corresponding 2016 – 2019 control periods. (crude rate ratio [CRR] 1.10, 95% CI 1.05 – 1.15, *P*<0.01). Using multivariable Poisson GLMM adjusted for age, sex and volume depletion, PE cases were significantly associated with the number of cases with pneumonia (CRR 1.09, 95 % CI 1.07−1.10, *P*<0.01), and with CTPA (CRR 1.10, 95 % CI 1.09−1.10, *P*<0.01). The increase of PE cases in 2020 compared with the control period remained significant (CRR 1.07, 95 % CI 1.02−1.12, *P*<0.01) when controlling for those factors. In the 2020 cohort, number of preceding average daily Covid-19 infections were associated with increased PE case incidence in all investigated windows, i.e. including preceding infections from 14 to 90 days. The best model (log likelihood −576) was with a window size of 4 days, i.e. average Covid-19 infections 14 – 17 days before PE hospitalization had a risk of 1.20 (95 % CI 1.12– 1.29, *P*<0.01).

**Conclusions:** There is an increase in PE cases since early May 2020 compared to corresponding periods in 2016 – 2019. This surplus was significant even when controlling for changes in potential modulators such as demographics, volume depletion, non-Covid-19 pneumonia, CTPA use and preceding Covid-19 infections. Future studies are needed (1) to investigate a potential causal link for increased risk of delayed PE with preceding SARS-CoV-2 infection, and (2) to define optimal screening for SARS-CoV-2 in patients presenting with pneumonia and PE.

---

After the first Covid-19 infection wave, we ^1^ and others ^2^ have observed a constant increase of pulmonary embolism (PE) hospitalizations not linked with active PCR-confirmed Covid-19. Compared with previous years, we found the 2020 surplus cohort to have less severe disease indicated by less thrombolytic therapy, intensive care, mechanical ventilation and in-hospital-mortality rates as well as shorter hospitalizations despite similar comorbidity burden.^1^

Potential contributors to this observation are unclear and could include undetected SARS-CoV-2 infection in pneumonia cases, more computed tomography use in suspected Covid-19 pneumonias, volume depletion due to the 2020 heat wave or complications of preceding Covid-19.^3–5^

Therefore, we analyzed associations between changes in PE hospitalizations and (1) the incidence of non-Covid-19 pneumonia, (2) the use of computed tomography pulmonary angiography (CTPA), (3) volume depletion and (4) preceding Covid-19 infection numbers in Germany.

Claims data of Helios hospitals in Germany were used and consecutive cases with a hospital admission between May 6 and December 15, 2020 (PE surplus period) ^1^ were analyzed and compared to corresponding periods covering the same weeks in 2016–2019 (control period). Crude rates for cases with (a) PE as target population (I26 as main discharge diagnosis according to ICD-10-GM), (b) pneumonia (J12 – J18), (c) volume depletion (E86) and (d) CTPA use (OPS 3-222 according to German procedure classification) were calculated by dividing the number of cumulative events by the number of days for each time period. Cases with PCR-confirmed SARS-CoV-2 infection (U07.1) were excluded. Crude rate ratios (CRR) were calculated using Poisson regression to model the number of hospitalizations.

We analyzed the number of cases in the target period with multivariable Poisson general linear mixed models (GLMM) including (a) cohorts of 2020 versus 2016–2019, (b) the number of cases with pneumonia, (c) CTPA, and (d) volume depletion and adjusted for age and sex.

In order to associate the daily number of PE cases in 2020 with the number of preceding SARS-CoV-2 infections in Germany,^6^ we calculated the average number of daily infections (divided by 10,000) occurring 14 up to 90 days with increasing window sizes before PE cases and modelled the data with Poisson regression.

We report CRR or odds ratios (OR, calculated by exponentiation of the regression coefficients) together with 95% confidence intervals (CI) for the comparisons of the different periods and *P* values for the interactions. For all tests, we apply a two-tailed 5% error criterion for significance.

This study was approved by the Ethics Committee at the Medical Faculty, Leipzig University (#490/20-ek). Due to the retrospective study of anonymized data, informed consent was not obtained.

There were 2,404 PE hospitalizations between May 6 and December 15, 2020 as opposed to 2,112 – 2,236 (total 8,717) in the corresponding 2016 – 2019 control periods. (CRR 1.10, 95% CI 1.05 – 1.15, *P*<0.01). As depicted in Figure 1, there were less pneumonias (CRR 0.85, 95% CI 0.83−0.86, *P*<0.01), more CTPA use (CRR 1.11, 95% CI 1.10−1.12, *P*<0.01) and less volume depletion (CRR 0.91, 95% CI 0.90−0.92, *P*<0.01) in the 2020 cohort. Compared with the control period, there was more frequent CTPA use in non-Covid-19 pneumonia cases without PE in 2020 (25.7 versus 19.1%, OR 1.37, 95 % CI 1.32–1.42, P<0.01). In cases with PE, pneumonias were more frequently observed in 2020 versus 2016−2019 (27.9 versus 25.1%, CRR 1.15, 95 CI 1.06−1.25, *P*<0.01). Vice versa, in cases with pneumonia, there were more cases with PE in 2020 versus 2016−2019 (5.3 versus 3.7%, CRR 1.44, 95 CI 1.35−1.55, *P*<0.01).

**Figure 1.**
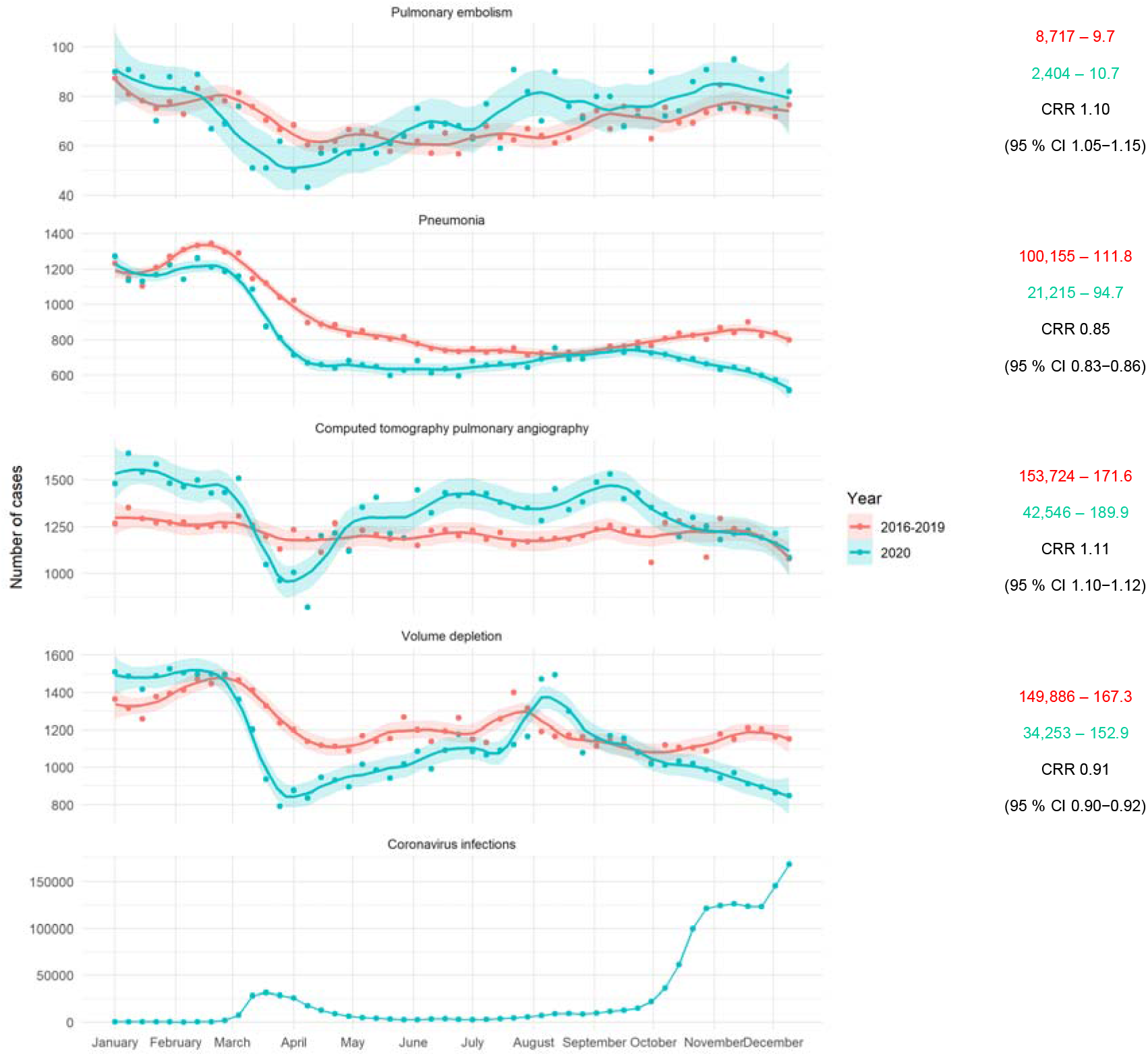
Left: Total weekly hospital admissions for pulmonary embolism, non-Covid-19 pneumonia, computed tomography pulmonary angiography, volume depletion and new SARS-CoV-2 infections in Germany (from top to bottom). Smooth curves for weekly admission rates were fitted via Locally Weighted Scatterplot Smoothing (LOESS) with a degree of smoothing of α = 0.2. Shaded areas represent 95% confidence intervals (CI). Right: Total and daily admissions, and resulting crude rate ratio (CRR) with 95% CI between May 6 and December 15, 2020 and the corresponding control period 2016 – 2019.

Using multivariable Poisson GLMM adjusted for age, sex and volume depletion, PE cases were significantly associated with the number of cases with pneumonia (CRR 1.09, 95 % CI 1.07−1.10, *P*<0.01), and with CTPA (CRR 1.10, 95 % CI 1.09−1.10, *P*<0.01). The increase of PE cases in 2020 compared with the control period remained significant (CRR 1.07, 95 % CI 1.02−1.12, *P*<0.01) when controlling for those factors.

In the 2020 cohort, number of preceding average daily Covid-19 infections were associated with increased PE case incidence in all investigated windows, i.e. including preceding infections from 14 to 90 days. The best model (log likelihood −576) was with a window size of 4 days, i.e. average Covid-19 infections 14 – 17 days before PE hospitalization had a risk of 1.20 (95 % CI 1.12– 1.29, *P*<0.01).

By analyzing claims data of the German-wide Helios hospital network, we have identified an increase in PE cases since early May 2020 compared to corresponding periods in 2016 – 2019.^1,2^ This surplus was significant even when controlling for changes in potential modulators such as demographics, volume depletion, non-Covid-19 pneumonia, CTPA use and preceding Covid-19 infections.

Pneumonia and PE are known to coexist ^7^ which is supported by the observed association between the two diseases. We could demonstrate that the total number of pneumonia cases was reduced in 2020, most likely due to social distancing and other lockdown measures, but at the same time CTPA use in general and in non-Covid-19 pneumonia cases (with even higher diagnostic sensitivity of pneumonia detection) increased. Those non-Covid-19 pneumonias could in fact be undetected SARS-CoV-2 infections with higher PE risk.^8^ Delayed PE following SARS-CoV-2 infection has also been reported,^3^ and PE was the reason for hospital readmissions in 0.6% patients after recovery from Covid-19 in previous studies.^4,5^ This observation is supported by our newly identified association between preceding Covid-19 and PE case numbers although this does not prove causality. Despite the inclusion of those variables into the multivariable prediction model, PE remained higher in 2020, suggesting that other undetected factors are contributing to this observation. For instance, use of glucocorticoids – frequently prescribed in SARS-COV-2 pneumonias – have been shown to associate with a higher PE risk within 30 days after exposure ^9^ but data on medication is not available in our cohorts. In addition, other factors such as hypercoagulability, platelet function and hypoxia which are associated with both Covid-19 infection and venous thrombosis including PE are also potential confounders but have not been analyzed.

Future studies are needed (1) to investigate a potential causal link for increased risk of delayed PE with preceding SARS-CoV-2 infection, and (2) to define optimal screening for SARS-CoV-2 in patients presenting with pneumonia and PE.

## Data Availability

. Helios Health and Helios Hospitals have strict rules regarding data sharing because of the fact that health claims data are a sensible data source and have ethical restrictions imposed due to concerns regarding privacy. Access to anonymized data that support the findings of this study are available on request from the Leipzig Heart Institute (www.leipzig-heart.de).

## Acknowledgments

DH was supported by the Volkswagen Foundation Germany through the Lichtenberg professorship program (# 84901).

